# Age-dependent Genetic Risk in Pulmonary Fibrosis Patients and Relatives

**DOI:** 10.64898/2026.01.07.25342496

**Authors:** David Zhang, Chad A. Newton, Imre Noth, Fernando J Martinez, Ganesh Raghu, Atlas Khan, Chen Wang, Matthew Moll, Michael H. Cho, Columbia Genomics Consortium, Krzysztof Kiryluk, Christine Kim Garcia

## Abstract

**Rationale:** Idiopathic pulmonary fibrosis (IPF) is an age-related disorder with common and rare genetic risk factors. It is unknown if the effects of PF genetic risk factors differ by chronologic age.

**Objectives:** To assess age-specific effects of genetic risk factors in PF patients and their relatives.

**Methods:** We identified common and rare genetic risk factors using a Columbia whole genome sequencing (WGS) cohort (777 IPF, 2905 controls) and replicated findings using Trans-Omics for Precision Medicine (TOPMed, 1148 IPF, 5202 controls). We assessed age-stratified genetic risk of IPF and assessed for interaction with age across a range of cutoffs. We analyzed 313 FPF pedigrees and compared age-specific prevalence of interstitial lung disease in relatives stratified by proband genetic risk factors.

**Measurements and Main Results:** Adjusted odds of disease from *MUC5B* SNP increase with age, while odds of disease from rare variants decrease with age. The magnitude of the interaction term between age and both genetic variables was greatest in younger individuals. There were significant interactions between age <55 and the *MUC5B* SNP (discovery p_interaction_=0.01; replication p_interaction_<0.0001) and rare variants (discovery p_interaction_<0.0001; replication p_interaction_=0.03). Pedigree analysis showed more prevalent disease especially in younger relatives in FPF families with rare variants versus without (p<0.0001).

**Conclusions:** Age modifies the effects of genetic risk factors in IPF. Rare variants confer greater risk in younger individuals whereas the *MUC5B* SNP confers greater risk in older individuals. Relatives of FPF patients with rare variants exhibit earlier prevalent disease, which has implications for preclinical disease screening.

## Introduction

Idiopathic pulmonary fibrosis (IPF) is the prototypical form of pulmonary fibrosis (PF) that carries a poor prognosis and has no cure^1^. Genetic studies of IPF have identified common and rare genetic risk factors associated with disease. A common *MUC5B* promoter polymorphism, present in ∼20% of European ancestry individuals, increases IPF risk 5-fold^2^. Polygenic scores of IPF and of telomere length derived from genome-wide association studies (GWAS) have been shown to independently increase disease risk^3–5^. Rare protein-altering variants in telomere biology genes (e.g. *TERT, TERC, PARN,* and *RTEL1*) and non-telomere genes (e.g. *SFTPC, SFTPA1/2,* and *KIF15*) increase disease risk 8- to 40-fold^3,6,7^. Cases with familial pulmonary fibrosis (FPF) are enriched 5-fold with rare disease-associated genes as compared to sporadic IPF^7^.

A conceptual framework for PF pathogenesis includes both inherited (genetic) and acquired causes (environmental factors and advancing age) in a “causal pie”^8^ that points to accelerated aging as a common underlying biologic pathway^9^. Multiple epidemiologic studies have demonstrated age-dependent increases in prevalence of IPF and other non-IPF interstitial lung disease (ILD)^10,11^. Interstitial lung abnormalities (ILA) represent potential preclinical precursors of PF and are similarly strongly associated with age^12^. Additionally, recent ATS society statements recommend screening relatives of FPF patients over the age of 50 for preclinical ILA, acknowledging its age-dependent nature^13^. Evidence in other age-related disorders demonstrate that genetic risk and age are not only component causes but may be interacting causes^14^. Higher polygenic risk of chronic obstructive pulmonary disease, heart disease, diabetes, atrial fibrillation, and certain cancers are associated with an earlier onset of disease^15,16^, suggesting that less acquired risk is necessary when a greater portion of causal risk is explained by genetic factors. Similarly, IPF patients with rare high-risk mutations have an earlier onset of disease^7,17^ – highlighting possible interacting risk. However, systematic assessment of interactions between age and IPF genetic risk factors have not been explored.

We perform the first study to formally assess interaction between age and IPF genetic risk factors and make the novel observation that age modifies the effects of the *MUC5B* SNP and rare variants in opposite directions. Rare variants confer greater IPF risk in younger individuals, while the *MUC5B* SNP confers greater risk in older individuals. We extend this finding to show that at-risk relatives of FPF patients with rare variants have earlier prevalent ILD. Our findings demonstrate the complex relationship between age and IPF genetic risk with implications for preclinical disease screening.

## Methods

Please see online supplement for detailed methods.

### Patient cohorts

The Columbia discovery IPF case-control cohort has been described previously^3,6,7^. The institutional review board at Columbia University Medical Center (AAAS0753, AAAS7495, and AAAP0052) approved this study. A replication cohort using data from the Trans-Omics for Precision medicine (TOPMed) program^18^ has been described previously^3,6^. In the TOPMed cohort, IPF individuals (phs001607) were included as cases and participants of the MESA (phs001416) and Framingham Heart (phs000974) studies were used as controls. Age of diagnosis was not available, so age at enrollment was used for all analyses.

### Genome sequencing, rare variants, and polygenic scores

Whole genome sequencing (WGS) data was generated for the Columbia cohort using standard protocols on Illumina’s NovaSeq 6000 platform. Raw sequencing reads were aligned to hg19 reference genome and cases and controls underwent joint genotyping according to GATK best practices. WGS data from the TOPMed cohort was aligned to hg38 reference genome and joint genotyped per published pipelines. All cohorts were pruned for relatedness within 3 degrees using KING^19^. Rare damaging missense or loss-of-function mutations in IPF-associated genes (*TERT, TERC, RTEL1, PARN, DKC1, NAF1, TINF2, SFTPC, SFTPA1/2, KIF15*) were identified using *in silico* tools as previously described^3,6,7,20^. Common variant genotyping and construction of polygenic scores was performed as previously described^3^. We applied a pruning and thresholding approach to include genome-wide significant risk variants identified from published GWAS studies of IPF and of telomere length. As described previously^3,4^, we assessed the *MUC5B* rs35705950 promoter polymorphism separately due to its outsized effect. We defined an IPF polygenic risk score using 12 SNPs^21^ excluding overlapping telomere-associated SNPs^22^ (*TERT* rs7725218, *TERC* rs12696304, *RTEL1* rs4130809) and the *MUC5B* SNP (IPF-PRS-M-T). We defined a telomere length polygenic score using 190 SNPs (TL-PRS). For sensitivity analysis we used alternative polygenic scores that “redistribute” the three shared loci, as well as an alternative IPF polygenic score using the lassosum method^4^ previously described.

### FPF Pedigrees

We leveraged a historic collection of FPF pedigrees to determine age of prevalent disease amongst affected and unaffected relatives of FPF patients. Pedigree construction was done using information collected from FPF probands identified through ILD clinic at UT Southwestern and Columbia University from 2005-2019. Probands for each pedigree have undergone WGS as previously described^6,7^ and have been analyzed for common and rare genetic risk factors. Medical information relating to case status for relatives were obtained from family members or directly by self-report. We abstracted characteristics of relatives including degree of kinship to proband, age, and presence or absence of fibrotic interstitial lung disease.

### Statistical Analysis

For primary analysis, we performed logistic regression to identify adjusted associations of genetic variables with IPF diagnosis stratified by age groups across decades of life. All analyses were adjusted for age, sex, and principal components of ancestry. We selected 5 principal components of ancestry based on visualization of variance explained on a scree plot as previously described^3^. To test for interaction, we assessed a range of age cutoffs from 50 to 80 and genetic variables by including both terms plus an interaction term in a multivariable model. Interaction beta coefficients and 95% confidence intervals were computed across all age cutoffs for both genetic variables and both cohorts. We reported the effect size of genetic variables on IPF risk dichotomized by age above/below 55 based on visualization of interaction term betas across the range of age cutoffs.

We also determined genetic makeup of IPF risk stratified by age group by utilizing the liability threshold model. This model transforms the observed probability of a binary trait to a continuous liability scale whereby exceeding a theoretical threshold results in development of that trait^23^. We compared the proportion of variance explained by common and rare genetic variables in full and reduced linear models removing the variable of interest. We then converted that proportion of variance to a liability scale accounting for ascertainment in case-control cohorts and assuming a fixed prevalence of IPF in the general population^24^.

For pedigree analysis, we visualized disease prevalences of 1^st^ and 2^nd^ degree relatives across age groups. We determined disease prevalences at each age group of relatives (# affected/total relatives) and estimated 95% confidence intervals using one sample proportion tests. We compared number of affected and unaffected relatives in each age group stratified by genetic risk factors of the affected proband in each family. We fit a generalized linear mixed-effects model (GLMM) with same covariates but including family as a random intercept to determine differences of the proportion of affected relatives.

## Results

### Age and genetic characteristics of cohorts

The Columbia discovery cohort included 777 unrelated IPF cases and 2905 controls and the TOPMed replication cohort included 1148 unrelated IPF and 5202 controls. Both Columbia and TOPMed IPF cohorts were predominantly of European ancestry (84% and 96%, respectively), while the control cohorts were more representative of non-European ancestry (27% and 46%, respectively). The median (IQR) age of enrollment for the Columbia and TOPMed IPF cohorts was 67 (61, 73) and 66 (60, 72) years, respectively. Separating by age groups (<50, 50-60, 60-70, 70-80, and >80), both discovery and replication IPF cohorts had similar age distributions (**Figure 1**). In both IPF cohorts, there were similar minor allele frequencies of the *MUC5B* SNP (Columbia MAF 0.34, TOPMed MAF 0.35) and proportions of rare variant carriers (Columbia carriers 12.1%, TOPMed carriers 12.7%).

**Figure 1.**
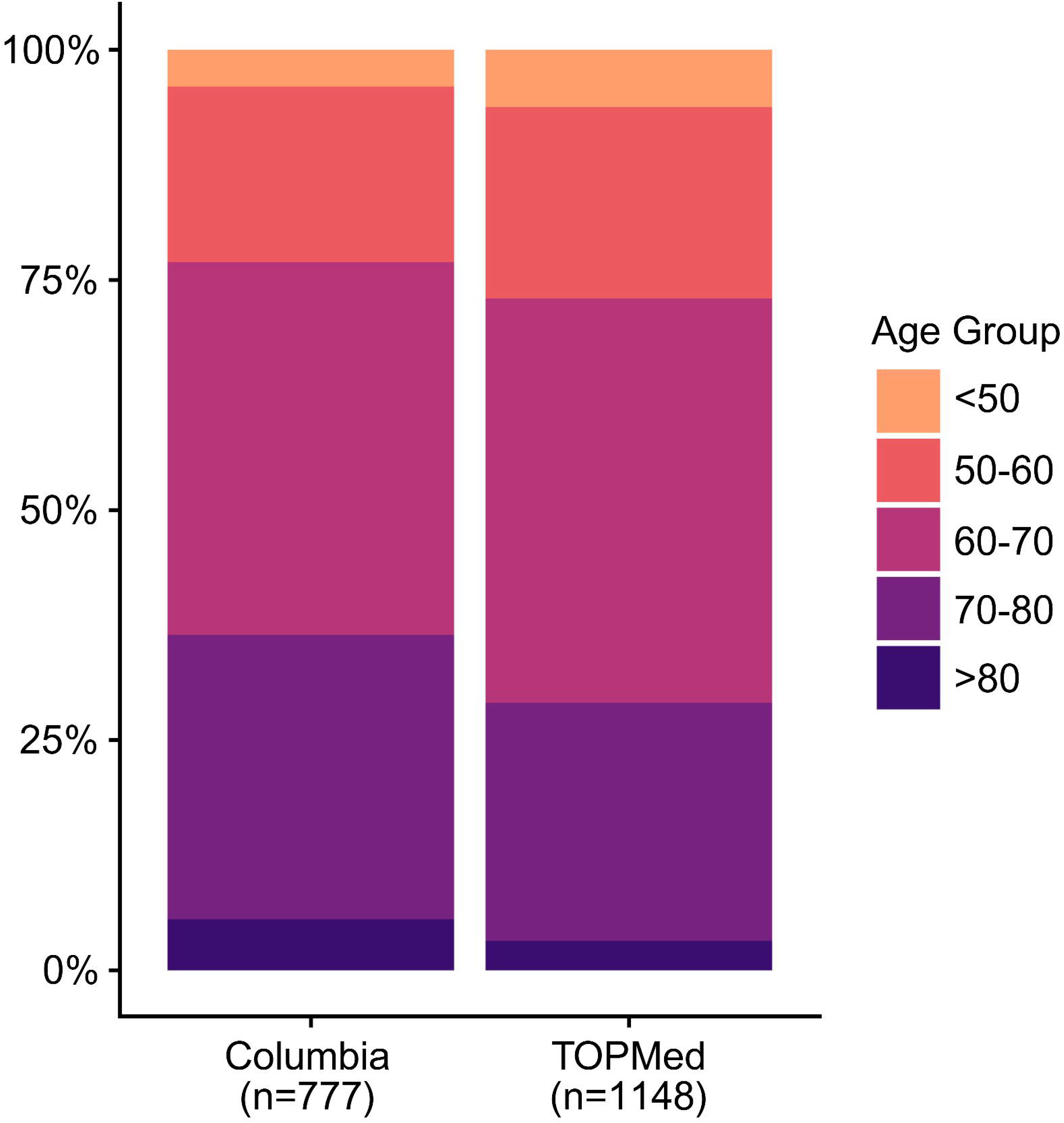
Age groups of IPF cohorts. Proportion of IPF subjects by age groups in the Columbia and TOPMed cohorts.

We observed notable differences of the genetic characteristics of IPF and controls across age groups for both cohorts (**Table 1**). Amongst controls, we found similar genetic characteristics across all age groups. In contrast, IPF rare variant carriers were consistently more prevalent amongst the <50 age group (39% in Columbia cohort, 27% in TOPMed cohort) and least prevalent amongst the >80 age group (5% in Columbia cohort, 3% in TOPMed cohort). Similarly, we observed the lowest minor allele frequency of the *MUC5B* SNP amongst IPF cases in the <50 age group (0.23 in Columbia cohort, 0.20 in TOPMed cohort) compared to the oldest age group (0.36 in Columbia cohort, 0.47 in TOPMed cohort). In contrast, there were inconsistent age group specific differences for both polygenic scores (IPF-PRS-M-T and TL-PRS).

**Table 1.** Age and genetic characteristics of case-control cohorts.

| Characteristics | Age <50 |  | Age 50-60 |  | Age 60-70 |  | Age 70-80 |  | Age >80 |  |
| --- | --- | --- | --- | --- | --- | --- | --- | --- | --- | --- |
|  | IPF | Control | IPF | Control | IPF | Control | IPF | Control | IPF | Control |
| **Columbia cohort** |  |  |  |  |  |  |  |  |  |  |
| Male, n (%) | 19 (61%) | 657 (54%) | 109 (74%) | 332 (59%) | 221 (70%) | 376 (61%) | 179 (75%) | 230 (62%) | 34 (79%) | 70 (51%) |
| Genetic ancestry, n (%) |  |  |  |  |  |  |  |  |  |  |
| European | 23 (74%) | 761 (63%) | 112 (76%) | 416 (73%) | 275 (87%) | 486 (79%) | 210 (88%) | 272 (73%) | 36 (84%) | 60 (44%) |
| Admixed American | 2 (6%) | 168 (14%) | 24 (16%) | 45 (8%) | 24 (8%) | 50 (8%) | 19 (8%) | 45 (12%) | 5 (12%) | 34 (25%) |
| African | 3 (10%) | 153 (13%) | 6 (4%) | 69 (12%) | 8 (3%) | 46 (7%) | 1 (2%) | 35 (9%) | 1 (2%) | 28 (21%) |
| East Asian | 1 (3%) | 57 (5%) | 3 (2%) | 22 (3%) | 1 (<1%) | 16 (3%) | 4 (2%) | 7 (2%) | 0 | 4 (3%) |
| South Asian | 1 (3%) | 36 (3%) | 1 (<1%) | 8 (1%) | 5 (2%) | 9 (1%) | 2 (<1%) | 6 (2%) | 0 | 1 (<1%) |
| Other | 1 (3%) | 41 (3%) | 2 (1%) | 6 (1%) | 2 (<1%) | 8 (1%) | 1 (<1%) | 7 (2%) | 1 (2%) | 9 (7%) |
| *MUC5B* SNP MAF | 0.23 | 0.11 | 0.32 | 0.14 | 0.34 | 0.12 | 0.37 | 0.16 | 0.36 | 0.15 |
| RV carriers, n (%) | 12 (39%) | 17 (1.4%) | 30 (20%) | 8 (1.4%) | 35 (11%) | 11 (1.8%) | 15 (6%) | 5 (1.3%) | 2 (5%) | 2 (1.5%) |
| IPF-PRS-M-T (mean±SD) | 0.13±0.75 | 0.00±0.99 | 0.61±1.1 | 0.01±0.96 | 0.54±1.0 | 0.00±1.05 | 0.59±1.0 | 0.05±1.02 | 0.42±1.1 | -0.14±0.91 |
| TL-PRS (mean±SD) | 0.41±0.75 | -0.02±0.99 | 0.52±1.1 | 0.00±0.96 | 0.45±1.0 | 0.05±1.05 | 0.44±1.0 | -0.03±1.02 | 0.23±1.1 | 0.00±0.91 |
| Total, n | 31 | 1216 | 148 | 556 | 315 | 615 | 240 | 372 | 43 | 136 |
| **TOPMed cohort** |  |  |  |  |  |  |  |  |  |  |
| Male, n (%) | 45 (63%) | 329 (47%) | 167 (70%) | 733 (49%) | 347 (69%) | 757 (47%) | 212 (71%) | 585 (51%) | 25 (68%) | 100 (42%) |
| Genetic ancestry, n (%) |  |  |  |  |  |  |  |  |  |  |
| European | 67 (94%) | 369 (53%) | 230 (96%) | 711 (47%) | 482 (96%) | 868 (53%) | 287 (97%) | 680 (59%) | 34 (92%) | 172 (73%) |
| Admixed American | 1 (1%) | 117 (17%) | 4 (2%) | 242 (16%) | 9 (2%) | 240 (15%) | 0 | 131 (11%) | 1 (3%) | 20 (8%) |
| African | 2 (3%) | 123 (18%) | 3 (1%) | 342 (23%) | 3 (<1%) | 332 (20%) | 1 (<1%) | 204 (18%) | 0 | 24 (10%) |
| East Asian | 0 | 76 (11%) | 1 (<1%) | 177 (10%) | 5 (1%) | 168 (10%) | 6 (2%) | 124 (11%) | 1 (3%) | 20 (8%) |
| South Asian | 1 (1%) | 0 | 0 | 0 | 1 (<1%) | 1 (<1%) | 2 (<1%) | 0 | 0 | 0 |
| Other | 0 | 11 (2%) | 1 (<1%) | 26 (2%) | 4 (<1%) | 16 (<1%) | 1 (<1%) | 8 (<1%) | 1 (3%) | 0 |
| *MUC5B* SNP, MAF | 0.20 | 0.08 | 0.31 | 0.08 | 0.36 | 0.07 | 0.38 | 0.07 | 0.47 | 0.09 |
| RV carriers, n (%) | 19 (27%) | 5 (0.7%) | 34 (14%) | 19 (1.3%) | 65 (13%) | 14 (0.8%) | 27 (9%) | 15 (1.3%) | 1 (3%) | 1 (0.4%) |
| IPF-PRS-M-T (mean±SD) | 0.61±0.89 | 0.01±0.99 | 0.63±0.91 | -0.07±1.00 | 0.52±1.0 | 0.01±1.00 | 0.64±0.87 | 0.05±0.98 | 0.54±0.99 | 0.10±1.03 |
| TL-PRS (mean±SD) | 0.21±0.89 | 0.07±0.99 | 0.37±0.91 | -0.02±1.00 | 0.44±1.0 | -0.02±1.00 | 0.33±0.87 | 0.00±0.98 | 0.48±0.99 | 0.07±1.03 |
| Total, n | 71 | 696 | 239 | 1498 | 504 | 1625 | 297 | 1147 | 37 | 236 |

Carriers of rare damaging variants in IPF-associated genes (*TERT*, *TERC*, *PARN*, *RTEL1*, *DKC1*, *TINF2*, *NAF1*, *SFTPC*, *SFTPA1/2*, *KIF15*).
Polygenic scores represented as control-normalized z-scores. IPF-PRS-M-T is computed excluding the *MUC5B* rs35705950 polymorphism and three overlapping telomere-associated loci. MAF, minor allele frequency

### Age-dependent genetic associations with IPF

We performed stratified analyses within each age group to find age-dependent genetic associations with IPF status (**Figure 2**). In both cohorts we identified stepwise *decrease* in adjusted odds of IPF from the *MUC5B* SNP with younger age. The youngest age group (<50) had the lowest odds of IPF from each copy of the *MUC5B* minor allele (OR_Columbia_ 3.27 [95% CI 1.69, 6.07], OR_TOPMed_ 1.89 [1.12, 3.13]), whereas the oldest age group (>80) had higher odds (OR_Columbia_ 4.35 [95% CI 1.93, 10.6], OR_TOPMed_ 11.2 [5.20, 27.2]). Conversely, we identified a stepwise *increase* in adjusted odds of IPF from carrying rare variants with younger age; the youngest age group had the *highest* odds of IPF from carrying rare variants (OR_Columbia_ 44.2 [17.9, 110], OR_TOPMed_ 68.8 [18.3, 417]). In individuals older than 80, carrying a rare variant had a non-significant association with IPF in both cohorts (OR_Columbia_ 9.45 [0.58, 219], OR_TOPMed_ 6.64 [0.25, 178]), with precision of estimates limited by sample size.

**Figure 2.**
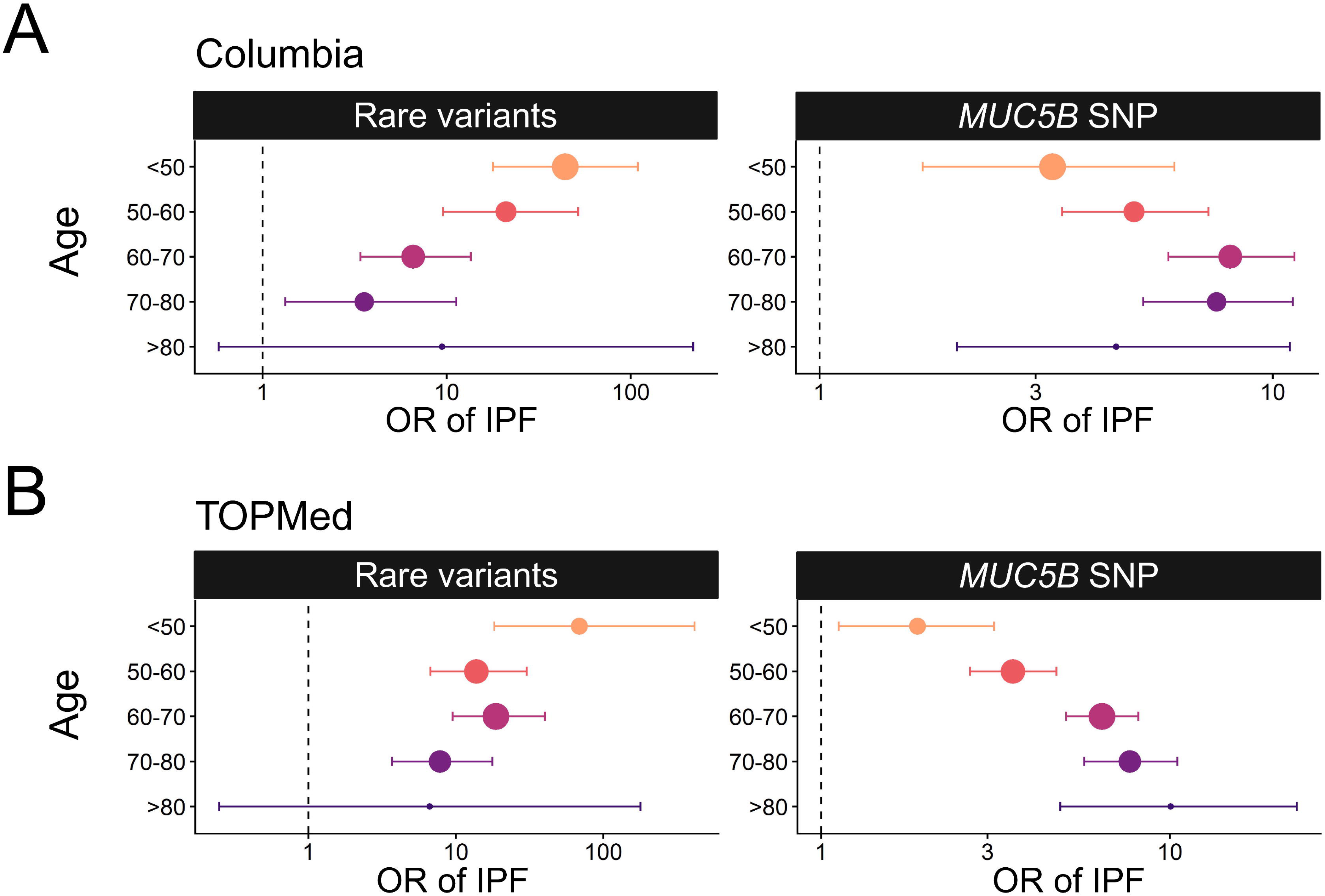
Genetic risk of IPF across age groups. Adjusted associations between the *MUC5B* rs35705950 polymorphism or rare variants and IPF risk stratified by age groups in **(A)** the Columbia discovery cohort and **(B)** the TOPMed replication cohort. Rare variants encompass damaging missense or loss-of-function variants in IPF-associated genes (*TERT, TERC, PARN, RTEL1, DKC1, TINF2, NAF1, SFTPC, SFTPA1/2, KIF15*). All associations adjusted for age, sex, and 5 principal components of ancestry.

In contrast, we did not observe a compelling trend across age groups for associations with both polygenic scores (IPF-PRS-M-T and TL-PRS) for either cohort (**Figure S1**). Alternative IPF polygenic scores also did not reveal age-dependent effects (**Figure S2**). To account for the possibility that the *MUC5B* trend was due to a relative depletion of that SNP in rare variant carriers, we performed sensitivity stratified analysis excluding rare variant carriers and found the same age-dependent associations (**Figure S3**).

### Interaction between age and IPF genetic variables

To confirm that genetic associations are age-dependent, we formally tested interaction between age and genetic variables on IPF risk across a range of age cutoffs between 50 and 80. We observed a consistent increase in the absolute magnitude of the beta coefficient of the interaction term with younger age cutoffs for rare variants and the *MUC5B* SNP, most evident with an age cutoff of 50-55 (**Figure 3A-B**). We then visualized the effect sizes of genetic variables dichotomized above or below 55 years of age (**Figure 3C-D**). In the Columbia discovery cohort, the *MUC5B* SNP conferred higher odds of disease in individuals >55 years old (OR 7.39 [6.02, 9.12]) than in those ≤55 years old (OR 3.81 [2.48, 5.87]; p_interaction_=0.01). In contrast rare variants conferred lower odds of disease in those >55 years old (OR 6.78 [4.16, 11.4]) compared to those ≤55 years old (OR 45.9 [20.5, 110]; p_interaction_=1.3×10^-5^). We identified similar interactions between age and genetic variables in the TOPMed replication cohort. The *MUC5B* SNP conferred higher odds of disease in older individuals (OR 6.14 [5.23, 7.23]) versus younger (OR 2.56 [1.82, 3.61]; p_interaction_=5.9×10^-6^) while rare variants conferred lower odds of disease in older individuals (OR 11.9 [7.70, 19.0]) versus younger (OR 35.5 [15.9, 88.5]; p_interaction_=0.03) in TOPMed.

**Figure 3.**
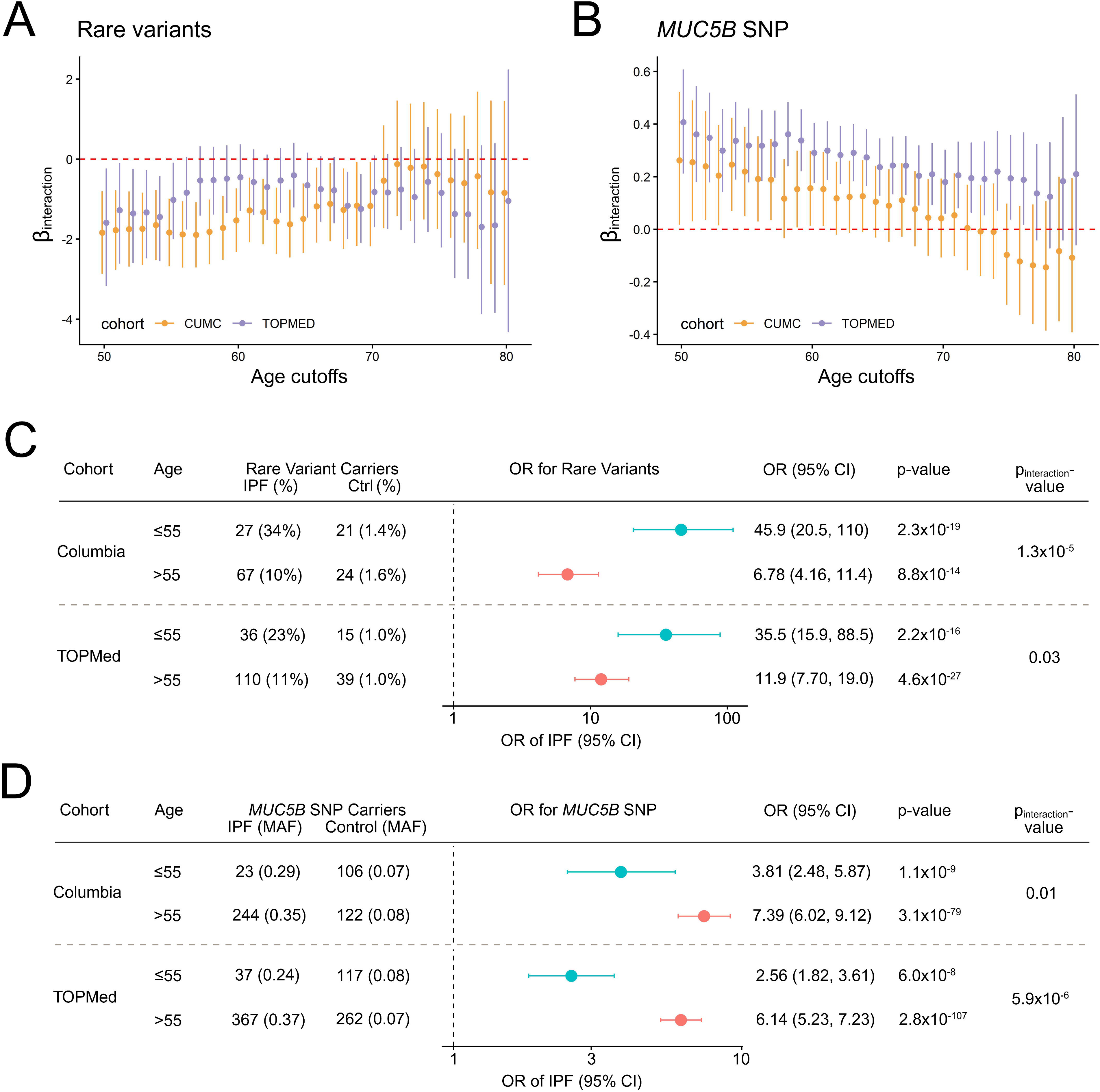
Interaction between age and key genetic risk factors in IPF. Visualization of beta coefficient and 95% confidence interval of interaction term between various age cutoffs and **(A)** rare variants or **(B)** *MUC5B* rs35705950 polymorphism and association with odds of IPF. Age cutoffs are assessed sequentially from 50-80 for both Columbia (orange) and TOPMed (purple) cohorts. Adjusted associations between **(C)** rare variants or **(D)** the *MUC5B* SNP and IPF risk stratified by age above or below 55 along with reported significance of interaction term. All associations adjusted for age, sex, and 5 principal components of ancestry. Abbreviations: MAF, minor allele frequency.

### Genetic makeup of young and old IPF patients

To visualize the genetic makeup of younger versus older IPF patients, we compared the contribution to genetic liability by known common and rare genetic risk factors across age bands (**Figure 4**). We find that across discovery and replication cohorts, there was a stepwise increase in genetic liability explained from rare variants with younger age and a stepwise increase in genetic liability explained from the *MUC5B* SNP with older age. In both cohorts, the youngest age group <50 had the most disease liability explained by genetic factors – the majority of which from rare variants. The genetic liability explained by RV in individuals <50 years old was greater in the Columbia cohort (45%) than in TOPMed cohort (15%).

**Figure 4.**
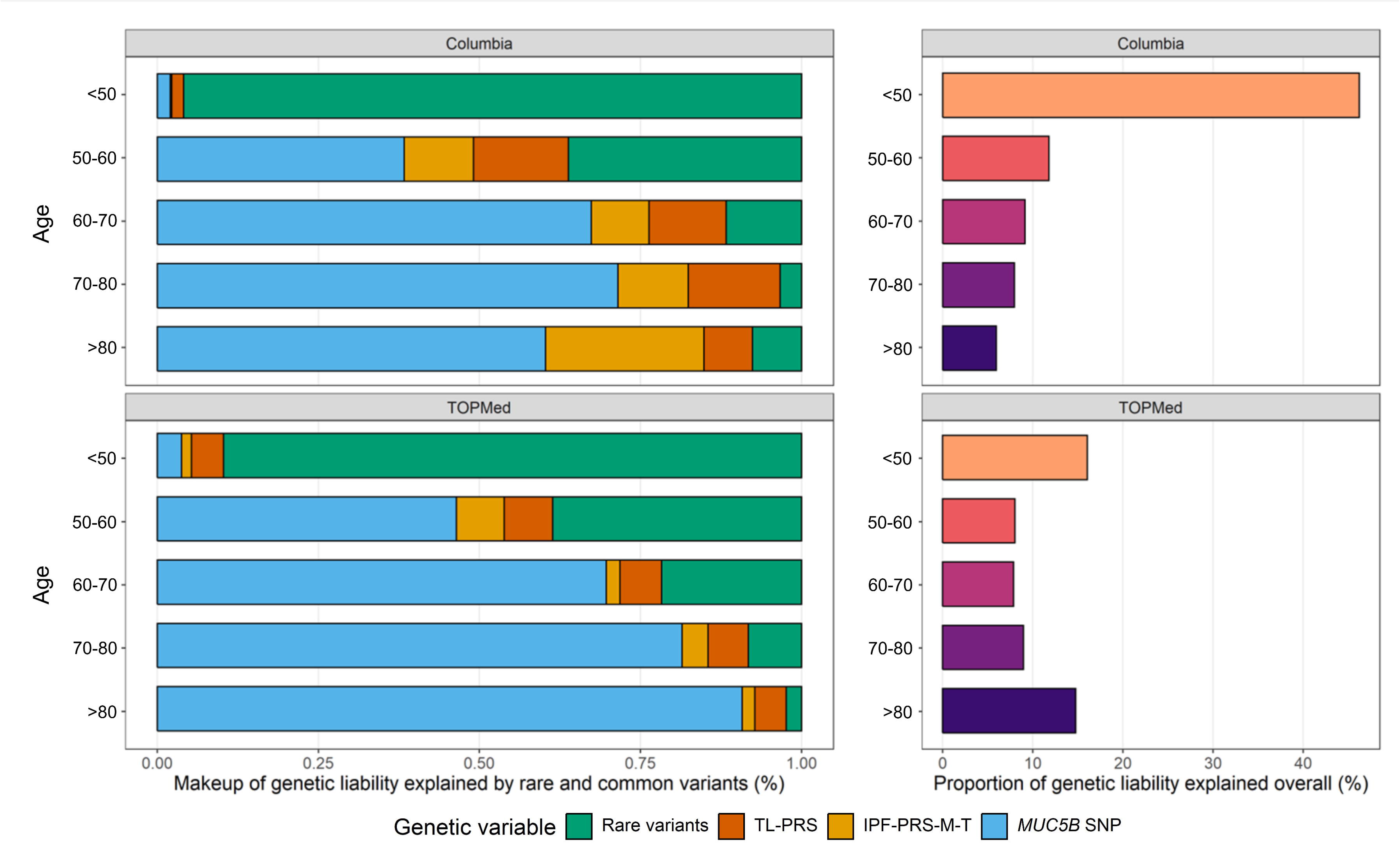
Genetic liability makeup of young versus old IPF patients. Contributions to overall genetic liability from known rare and common variants across age groups of IPF. Across both cohorts, younger patients have increased genetic liability explained by rare variants, while older patients have increased genetic liability explained by the *MUC5B* rs35705950 polymorphism. Overall genetic liability explained is highest in IPF subjects under 50 years old, making this the most genetically enriched group.

### Age-dependent disease prevalence in FPF relatives based on proband genetic factors

To determine age-dependent genetic risk in at-risk FPF relatives, we analyzed 313 FPF pedigrees with (n=95) and without (n=218) rare damaging variants in the proband. In total we included 2,539 first- and second-degree relatives. Rare variants were found in *TERT* (n=51), *TERC* (n=8), *RTEL1* (n=12), *PARN* (n=9), *KIF15* (n=6), *SFTPC* (n=5), *NAF1* (n=2), *DKC1* (n=1), and *SFTPA2* (n=1). Overall, we found a higher ILD prevalence in relatives of FPF patients with a rare variant (24% vs 18%, p_GLMM_=3.6×10^-6^ **Figure 5**). In stratified analysis, we find that the greatest difference in ILD prevalence occurred in younger age relatives aged 40-50 (18% vs. 5%, p_GLMM_=0.006) and aged 50-60 (28% vs. 14%, p_GLMM_=0.002). In families where a rare variant was identified in the proband, 69/179 (39%) of affected relatives were below age 60. In contrast, in families where no rare variant was identified, only 64/321 (20%) affected relatives were below age 60.

**Figure 5.**
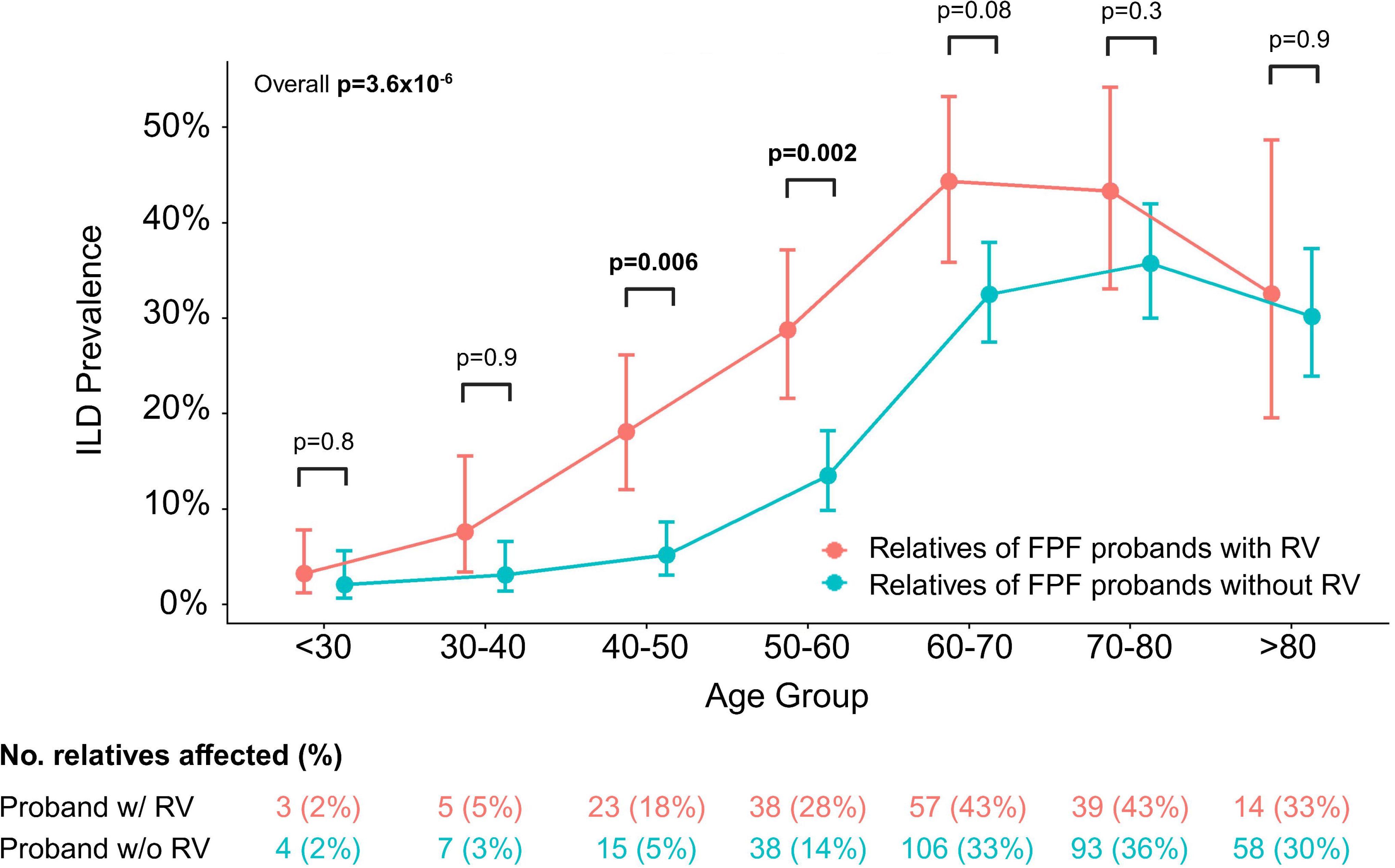
Prevalence of interstitial lung disease in relatives of FPF patients with or without rare variant. Prevalence of ILD amongst 1^st^ and 2^nd^-degree relatives of FPF patients across age groups from 313 FPF pedigrees. Relatives are stratified by if the proband was found to have a rare variant (red) or not (blue). P-values from generalized linear mixed-model (GLMM) are shown stratified by each age group, or overall adjusting for age. All associations are adjusted for by age and sex as fixed effects and family unit as a random effect.

For sensitivity analysis we separated FPF relatives of *TERT* carriers vs non-*TERT* carriers and found consistent trends (**Figure S5**). We also stratified relatives by proband *MUC5B* SNP status and by high (quartile 1) vs low (quartile 2-4) polygenic risk for both IPF (IPF-PRS-M-T) and telomere length (TL-PRS). We did not observe a statistically significant difference in disease prevalence in relatives when applying these classifications (**Figure S6**).

## Discussion

Our study leveraged two large cohorts of pulmonary fibrosis patients to characterize the interaction between genetic risk and age. We find a robust and reproducible gene-by-age interaction for the *MUC5B* common risk variant and rare variants in opposite directions, suggesting that the genetic makeup of younger versus older IPF patients is quantifiably different. We show that ILD prevalence is higher amongst younger (age 40-60) relatives of FPF patients with rare damaging variants compared to those without mutations. Since it can take up to a decade for early interstitial lung abnormalities to transform into definite fibrosis on CT, screening at age 50 may miss early ILA in two out of five relatives of FPF patients with rare damaging variants.

Many age-related disorders feature complex relationships between age and genetic risk. In a conceptual model where both genetic and acquired factors confer additive disease risk, having more genetic risk factors means that less acquired risk is necessary to develop disease, thereby leading to earlier disease onset. We confirm this phenomenon in IPF, where carrying a rare disease-associated mutation results in earlier age of diagnosis^7,17^ and confers higher risk at a younger age. This non-uniform effect of rare variants across age leads to a diminished and non-significant association with IPF risk in individuals over 80. We surmise that rare variant carriers who develop later onset of disease or are disease-free may have less environmental injuries or protective genetic or non-genetic factors that modify the effects of these high-risk mutations.

Interestingly, we identify a non-uniform association across age in the opposite direction with the *MUC5B* SNP which confers greatest genetic risk of IPF in the oldest age group. We find that this effect persists even when excluding rare variant carriers known to have decreased *MUC5B* minor allele frequencies^7,25,26^. This unexpected association mirrors prior reports of *MUC5B* minor allele carriers having older age of IPF diagnosis^27^ and suggests that it is not a typical genetic risk factor. The known dual role of the *MUC5B* SNP as a protective factor against respiratory infections^28^ (*e.g.* COVID-19^29–31)^ but a risk factor for pulmonary fibrosis may lead to complex associations with survival. The association between the *MUC5B* SNP and improved survival for IPF has been described^27,32^, but it remains unclear if index case bias is responsible^33,34^. Post-COVID survivors, but not decedents, with pulmonary fibrosis also feature increased *SCGB1A1* and *MUC5B* co-expressing cells in pulmonary terminal bronchioles in the absence of the *MUC5B* risk allele^35^. Together, these effects may contribute to a phenomenon whereby IPF patients with the *MUC5B* SNP are identified later in life.

We do not identify a consistent age-dependent effect for either the IPF or telomere-related polygenic risk factors. This contrasts with published findings in common diseases such as coronary disease, atrial fibrillation, diabetes, and cancer where higher polygenic risk leads to greater disease risk in younger age groups^15,36^. An accumulation of acquired non-genetic risk factors, including environmental exposures or comorbid conditions, may “dilute” the effect of polygenic risk with older age, rendering an attenuated effect. Interestingly, IPF rare variants – not common variants – behave in a similar fashion with decreased effect in older age. Polygenic risk may act differently in IPF either due to additional roles as genetic modifiers^3^ or due to limitations of underpowered GWAS studies to identify highly predictive scores. Furthermore, the age distribution of the discovery GWAS study plays a critical role in PRS predictive accuracy of age-related disorders in external cohorts^14^. Our findings demonstrate that subject age is a major source of heterogeneity of genetic makeup in IPF. Carefully balanced representation of IPF age groups in GWAS cohorts may be necessary for improving polygenic-based prediction.

Recognition of the differences in IPF genetic risk by age will have important implications for screening practices, especially amongst relatives of FPF patients. About 25% of FPF patients carry high-risk rare variants^7,37^. We find that that relatives of FPF rare variant carriers have higher disease prevalence, particularly in the 40 to 60-year age group, where 20-30% are affected by ILD. Affected relatives of rare variant carriers are also likely to carry the same high-risk rare variant themselves^38^, leading to rapid progression and early mortality or need for transplant^7,17,26,39^. Since the evolution from preclinical ILA to definite pulmonary fibrosis can take over a decade^40^, these relatives are particularly vulnerable to being missed by a screening cutoff age of 50 per recent ATS statements^13^. Our findings make the case for precision screening of preclinical ILA in FPF relatives by employing guideline-recommended^41,42^ genetic screening of the affected proband. Identification of a pathogenic variant in the proband should prompt earlier screening of at-risk relatives, although additional studies are needed to determine the optimal age cutoff for screening.

Our study has several limitations. Age of enrollment, not age of diagnosis, was available in all cohorts. Given the median survival of IPF of 3-5 years^1^, we surmise that enrollment age can serve as a surrogate for age of diagnosis. We additionally bin our cohorts into decades of age to attenuate the effects of misclassification. Due to inconsistent clinical phenotyping between cohorts, relevant environmental risk factors such as smoking or other environmental exposures were not available for study. Future studies that can incorporate comprehensive clinical risk along with genetic risk can further refine this gene-by-age effect^43^. While rare IPF mutations have similar prevalences and effects across genetic ancestry groups^44,45^, common risk variants like the *MUC5B* SNP are ancestry-dependent^46^; it remains unknown if these interactions are relevant across non-European ancestries – our data are underpowered to answer these questions.

Retrospective pedigree analysis was used to estimate disease prevalence amongst FPF relatives. Recall or observer bias may exist, and future prospective at-risk studies are needed to validate our findings. Nevertheless, our findings are in line with prior studies demonstrating greater disease prevalence in FPF families with pathogenic genetic risk factors^17^.

In conclusion, age and genetic risk factors are component causes and interacting causes for pulmonary fibrosis. Rare variants confer greater genetic risk of IPF in younger individuals, while the *MUC5B* SNP confers greater genetic risk of IPF in older individuals. This quantifiable difference in IPF genetic makeup across age groups is particularly relevant for FPF families with rare variants, where two out of five affected relatives with established ILD are below age 60 and may be screened too late for early intervention.

## Data Availability

Genotype data for discovery cohort allowable under consent is available in the Database of Genotypes and Phenotypes (dbGAP) under the following projects: Pulmonary Fibrosis and Telomerase Dysfunction, phs002692; Genomics of Glomerular Disorders, phs002480; Genomic Translation for ALS Care (GTAC), phs02973. The TOPMed cohort genetic and phenotypic data are available under dbGAP (Multi-ethnic Study of Atherosclerosis, phs0001416; Framingham Heart Study, phs000974; Idiopathic Pulmonary Fibrosis, phs001607).

## Acknowledgements

The authors thank the research participants for their participation and enrollment in study cohorts. The authors thank the investigators and institutions who supported the NHLBI TOPMed cohorts (IPF, MESA, and the Framingham Heart Study). The authors would like to acknowledge the Collaborative Group of Genetic Studies of IPF for access to summary statistics of prior IPF GWAS. The <u>Columbia Genomics Consortium</u> includes genomic data from a number of projects and contact investigators: the Columbia University Biobank (Muredach Reilly), the Columbia CKD Biobank (Krzysztof Kiryluk and Ali Gharavi), Genomic Translation for ALS Care (GTAC; Matthew Harms), Pulmonary Fibrosis (Christine Garcia), Atopic disease (Joshua Milner) and Prenatal disease (Ronald Wapner). Only the contact investigator(s) for each project are listed.

