## Supplementary material for "Age-dependent Genetic Risk in Pulmonary Fibrosis Patients and Relatives": online supplement

### Methods

**Figure S1. Age-stratified IPF-associated and telomere-related polygenic risk**

**Figure S2. Age-stratified IPF-associated polygenic risk using alternative score**

**Figure S3. Age-stratified common genetic risk of IPF excluding carriers of rare variants.**

**Figure S4. Prevalence of ILD in at-risk relative stratified by proband *TERT* and non-*TERT* rare variants**

**Figure S5. Prevalence of ILD in at-risk relative stratified by proband genetic status.**

**Methods**

*Patient cohorts*

The discovery IPF cohort sequenced at Columbia University has been described previously. Patients were enrolled from Columbia University Medical Center (CUMC), the University of Texas Southwestern, and the IPF Clinical Research Network PANTHER-IPF[3] (Evaluating the Effectiveness of Prednisone, Azathioprine, and N-acetylcysteine in Patients With IPF; NCT00650091) and ACE-IPF[4] (Anticoagulant Effectiveness in Idiopathic Pulmonary Fibrosis; NCT 00957242) clinical trials including only those individuals who consented to participate in both the parent study and the optional genetic substudy. Patients with IPF diagnosis meeting current guidelines were included in this study. Non-IPF disease controls were used for the discovery cohort as previously described.  Genome sequencing data from the TOPMed replication cohort was accessed through the database of Genotypes and Phenotypes (dbGaP) with permission. Genotype data for discovery cohort allowable under consent is available in the Database of Genotypes and Phenotypes (dbGAP) under the following projects: Pulmonary Fibrosis and Telomerase Dysfunction, phs002692; Genomics of Glomerular Disorders, phs002480; Genomic Translation for ALS Care (GTAC), phs02973. The TOPMed cohort genetic and phenotypic data are available under dbGAP (Multi-ethnic Study of Atherosclerosis, phs0001416; Framingham Heart Study, phs000974; Idiopathic Pulmonary Fibrosis, phs001607).

*Genomic data acquisition and processing*

Whole genome sequencing was performed for the discovery cohort at Columbia University using Illumina's NovaSeq 6000 platform with standard protocols as previously described. Raw sequencing reads were aligned to GrCh37 using DRAGEN and sample-level BAM files were jointly genotyped per GATK best practices. Site-level QC was performed to exclude variants failing variant quality score recalibration (VQSR) filters. Variant annotation was performed using Variant Effect Predictor (VEP[1]) and dbNSFP[2]. Rare qualifying variants in IPF-associated genes were identified applying gnomAD[3] population-specific allele frequencies below 0.0005 and in silico prediction of loss of function or missense mutations passing consensus predictors (Polyphen2[4] probably damaging, REVEL[5] score ≥ 0.5, and PrimateAI[6] score ≥ 0.8). *TERC* non-coding variants were included if population allele frequencies were below 0.0005 and disrupted intramolecular base-pairing. Principal components of ancestry were obtained using *plink*[7] after pruning variants for linkage disequilibrium.

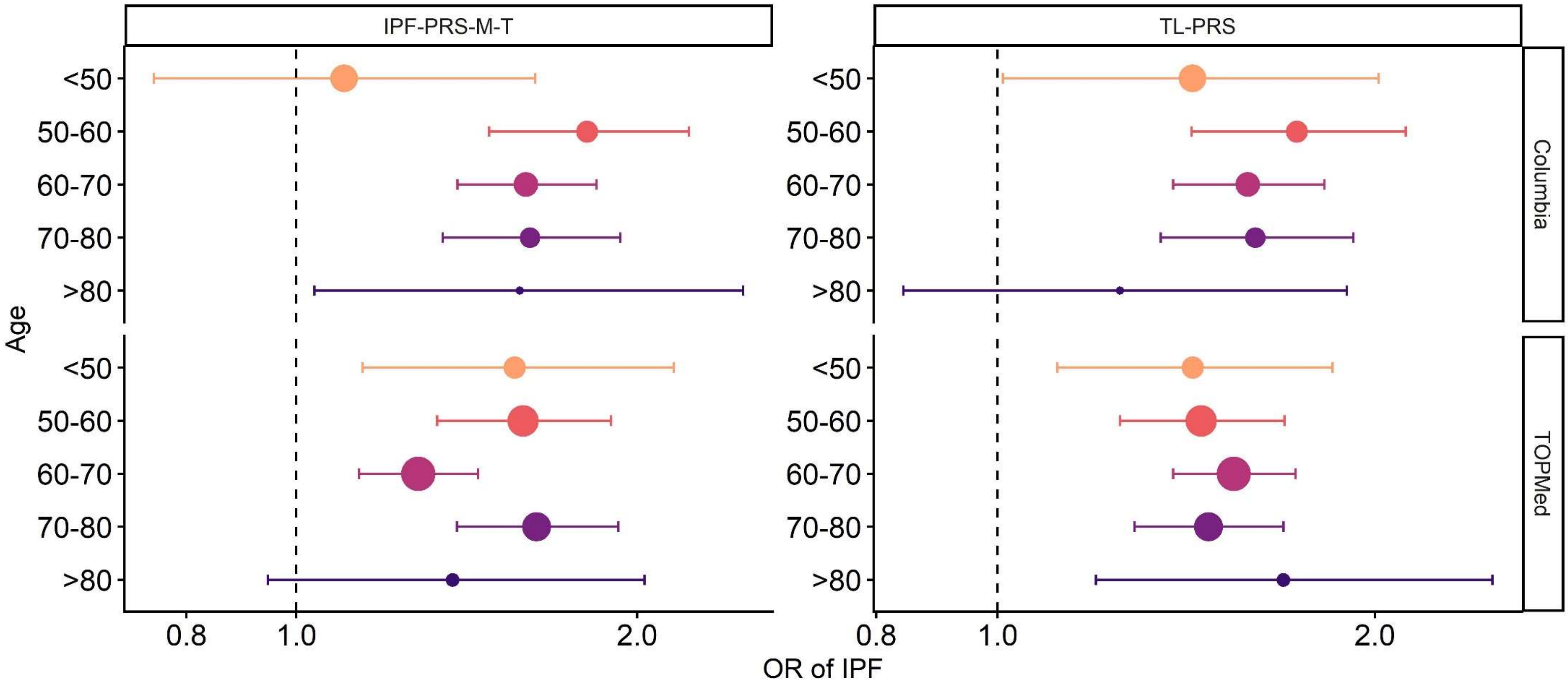


**Figure S1. Age-stratified IPF-associated and telomere-related polygenic risk.** Stratified associations by age group show inconsistent age-dependent effects in both cohorts. The IPF-PRS-M-T comprises 12 SNPs associated with IPF and excludes the *MUC5B* rs35705950 SNP and three telomere-associated loci (*TERT* rs7725218, *TERC* rs12696304, *RTEL1* rs41308092). All associations adjusted for age, sex and 5 PC of ancestry.

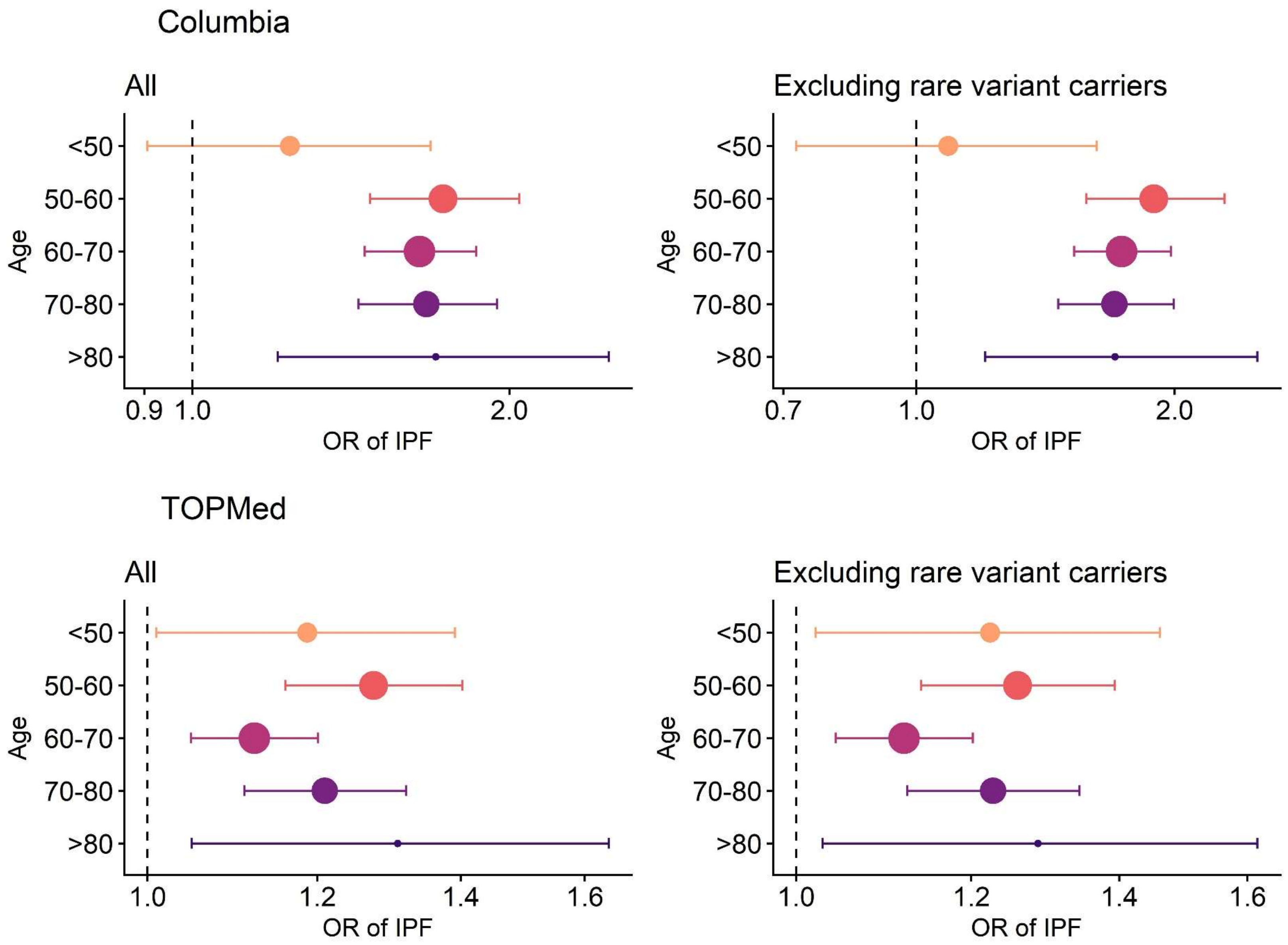


**Figure S2. Age-stratified IPF-associated polygenic risk using alternative score.** Alternative IPF polygenic score using the lassosum method excluding a 500kb region flanking the *MUC5B* polymorphism as previously described[8]. Stratified associations by age group show inconsistent age-dependent effects across cohorts before and after excluding rare variant carriers. In the Columbia cohort, the polygenic score is specifically not associated in those age below 50, whereas the score is similarly associated across age groups in TOPMed. All associations adjusted for age, sex and 5 PC of ancestry.

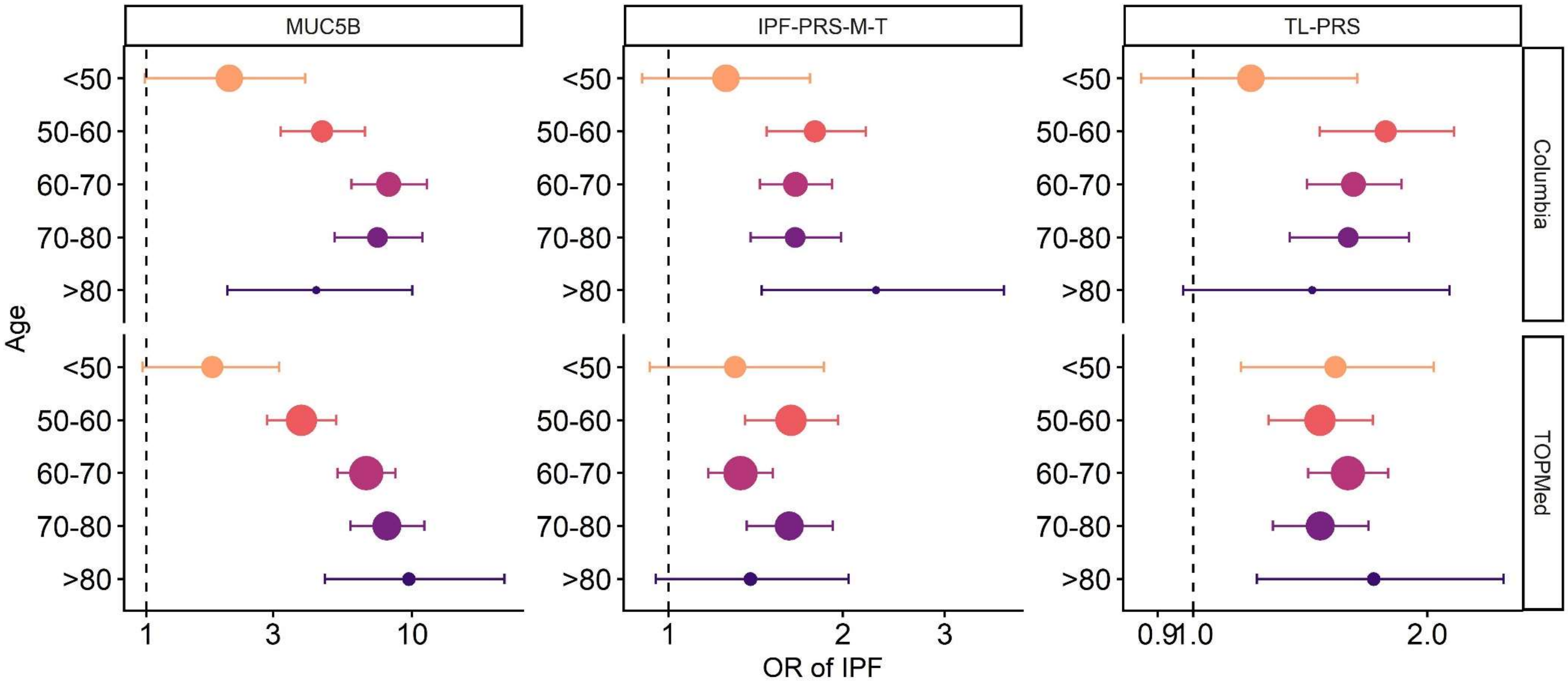


**Figure S3. Age-stratified common genetic risk of IPF excluding carriers of rare variants.** Stratified associations by age group show inconsistent age-dependent effects in both cohorts after excluding rare variant carriers. The IPF-PRS-M-T comprises 12 SNPs associated with IPF and excludes the *MUC5B* rs35705950 SNP and three telomere-associated loci (*TERT* rs7725218, *TERC* rs12696304, *RTEL1* rs41308092). All associations adjusted for age, sex and 5 PC of ancestry.

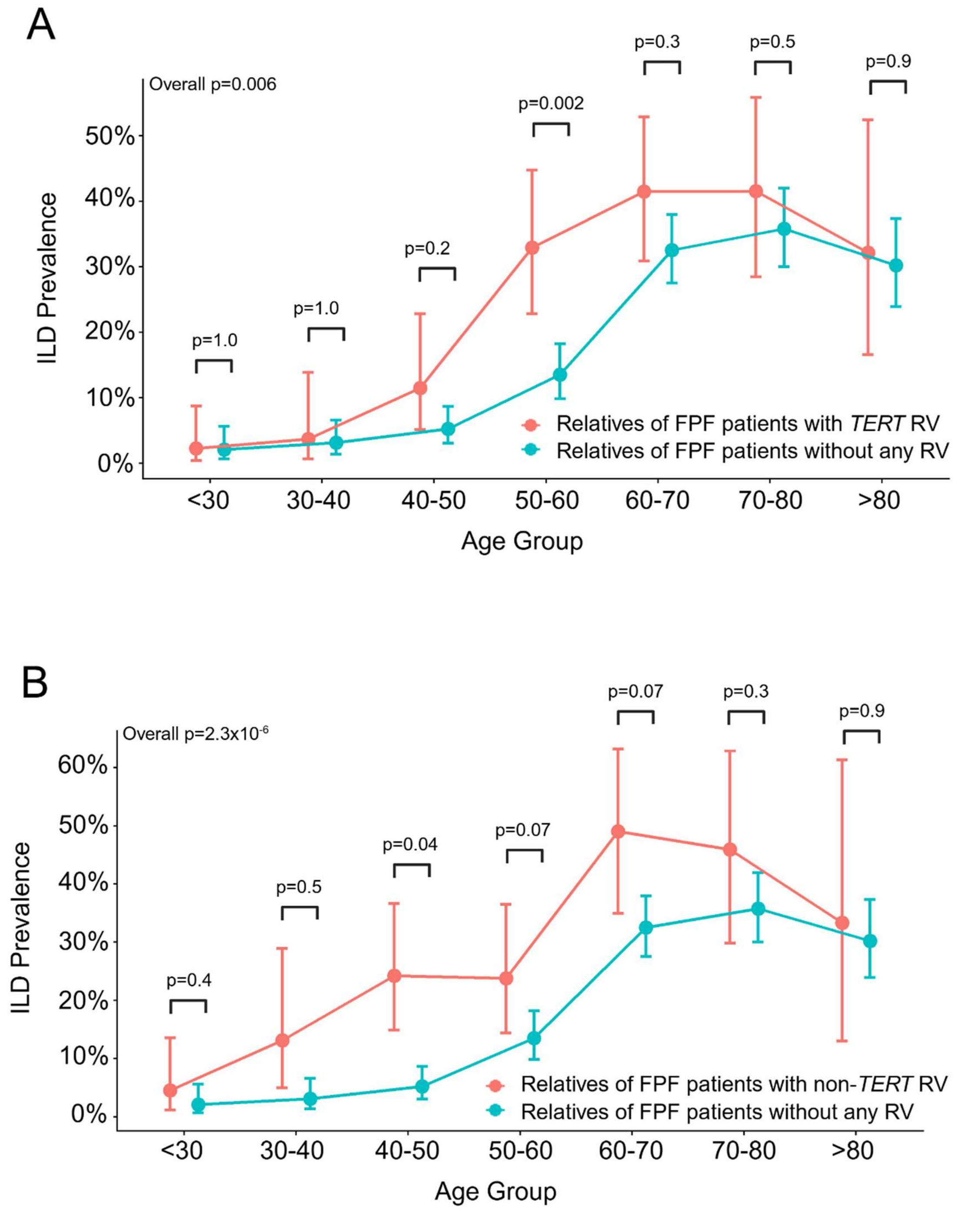


**Figure S4. Prevalence of ILD in at-risk relative stratified by proband *TERT* and non-*TERT* rare variants.** Visualization of raw ILD prevalences in 1$^{st}$ and 2$^{nd}$ degree relatives of FPF patients stratified by genetic risk factor across age groups. Relatives are stratified into FPF families with no rare variants (blue) or **(A)** *TERT* rare variants (red) and **(B)** non-*TERT* rare variants (red). There were significant overall differences in ILD prevalences amongst relatives of FPF patients with no rare variants vs. *TERT* rare variants and vs. non-*TERT* rare variants.

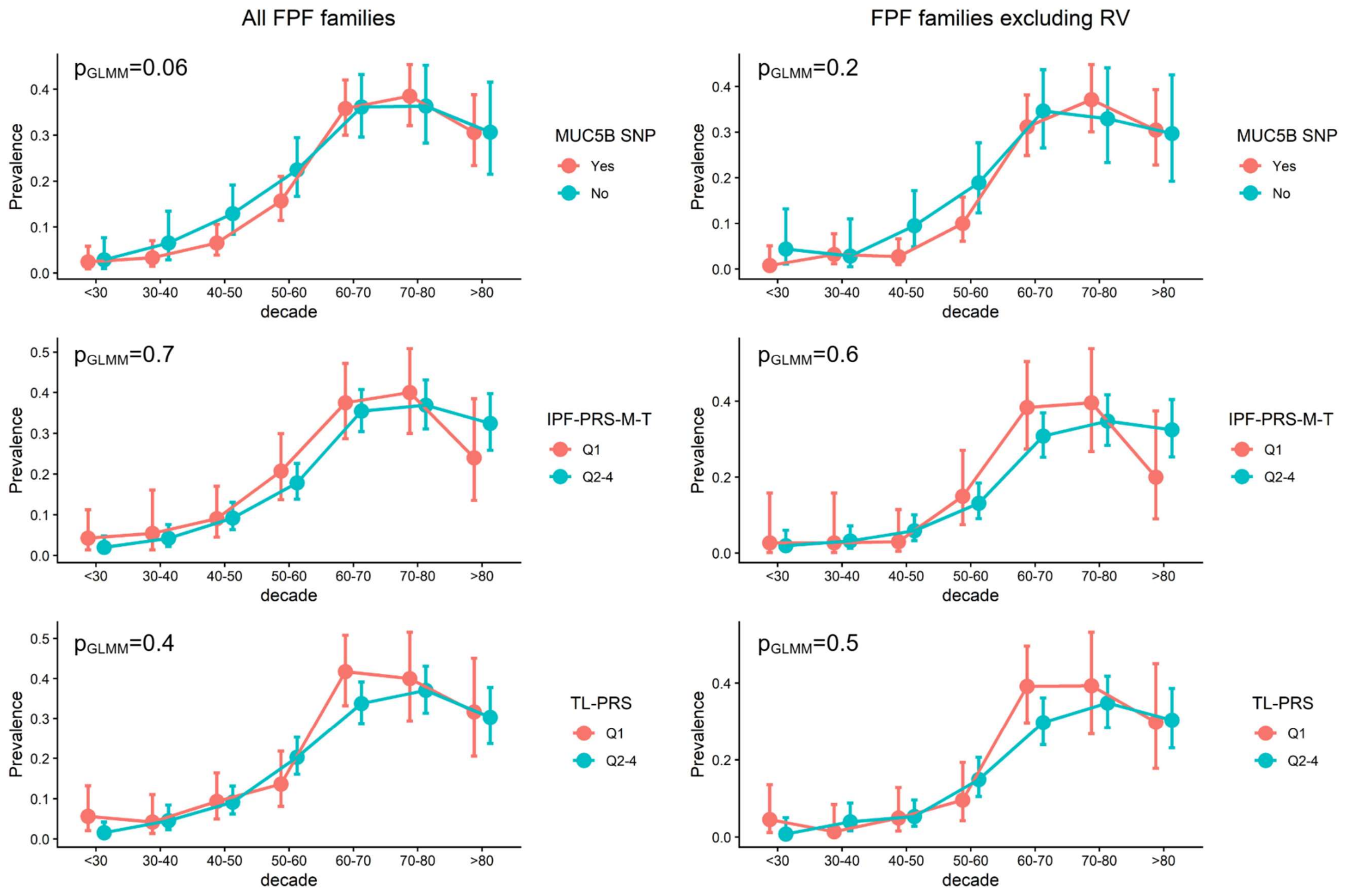


**Figure S5. Prevalence of ILD in at-risk relative stratified by proband genetic status.** Visualization of raw ILD prevalences in 1st and 2nd degree relatives of FPF patients stratified by genetic risk factor across age groups. Relatives are stratified into FPF families with a higher genetic risk (red) or lower (blue). Prevalences are shown both inclusive and exclusive of families where a rare variant was identified in the proband. No stratification scheme showed significant differences in prevalence.
